# Computational Phenotypes for Temporomandibular and Orofacial Pain Disorders

**DOI:** 10.64898/2026.09.04.26351324

**Authors:** Glenn Thomas Clark, Anette Vistoso Monreal, Nicolas Veas, Jianfu Chen, Gerald E. Loeb

## Abstract

**Background:** Temporomandibular disorders (TMDs) and orofacial pain (OFP) conditions affect approximately one-third of the global population, yet diagnosis often relies on subjective clinical assessment rather than standardized, evidence-based criteria. This diagnostic uncertainty contributes to misdiagnosis, misdirected or delayed treatment and significant healthcare costs.

**Case description:** We prospectively collected structured clinical data from 1,501 patients using a custom, structured, note-documentation system designed for machine learning compatibility (Smart Medical Note [SmartNote]). From the 126 possible TMD-OFP diagnoses, we identified 15 conditions with sufficient case volumes (≥24 exemplars) for analysis. The clinical features present in>50% of these cases underwent Monte Carlo statistical analysis to determine which are most useful for creating an objective diagnostic profile. The 15 evidence-based diagnostic profiles demonstrated strong discriminative performance (median AUC values 0.64-0.96).

**Practical Implications:** These diagnostic profiles represent the first large-scale, statistically derived criteria for TMD and orofacial pain conditions. By providing objective, quantifiable diagnostic standards, this statistical analysis of structured electronic records can support algorithmic diagnostic-assist software, reduce the time to appropriate treatment, and enhance clinical decision-making and documentation of diagnoses for dental practitioners, regardless of their level of specialized training in orofacial pain. Diagnostic alignment analysis confirms complete concordance with DC-TMD criteria for seven major diagnostic categories, validating the clinical applicability of our data-driven approach.

## Introduction

Temporomandibular disorders and orofacial pain conditions represent a diagnostically challenging area in dental practice. Recent comprehensive studies indicate that TMDs affect 31-34% of the global population, making them the second most common musculoskeletal condition after chronic low back pain, with an annual healthcare burden exceeding $4 billion in the United States alone [1,2]. Despite this prevalence, these conditions remain diagnostically difficult because of the typical absence of objective, standardized diagnostic criteria (e.g. serologic, histopathologic, imaging or any genetic markers). The full diagnostic landscape spans three major domains: musculoskeletal conditions (e.g., masticatory myalgia, arthralgia, disc displacements, osteoarthritis), parafunctional and behavioral conditions (e.g., bruxism, clenching, medication-induced clenching), and neuropathic/neurovascular conditions (e.g., trigeminal neuralgia, burning mouth syndrome, dysesthesia, tinnitus). All three domains are represented in this study’s 15 analyzed conditions.

### The Current Diagnostic Challenge

The complexity of TMD and OFP diagnosis stems from several factors. With over 77 different diagnostic categories, overlapping symptom profiles are common, and many patients present with multiple conditions simultaneously [3]. Current diagnostic approaches rely heavily on subjective clinical interview and examination-based assessments, at least some of which may be overlooked, ambiguously recorded or unrecorded in the free-text narrative note. Currently, diagnosis is based largely on expert opinion, leading to significant variability in diagnosis and treatment recommendations among practitioners.

It is important to emphasize that expert-derived diagnostic frameworks have played an essential role in the clinical management of TMD and orofacial pain conditions, particularly in the absence of objective biomarkers. Expert clinical judgment remains indispensable at the point of care. However, expert-opinion–based systems are inherently difficult to test, quantify, reproduce, and systematically refine, especially across diverse clinical settings and practitioner experience levels. These limitations motivate the development of complementary, data-driven approaches that can formalize, evaluate, and continuously update expert knowledge using real-world clinical data.

This diagnostic uncertainty has real consequences. Studies document average diagnostic delays of 3-7 years for conditions such as burning mouth syndrome and persistent facial pain, during which patients often undergo unnecessary dental procedures [4]. For dental practitioners, the lack of standardized criteria can lead to diagnostic uncertainty, inappropriate referrals, and misapplied treatment concerns.

### A Data-Driven Solution

Recent advances in artificial intelligence and machine learning present significant opportunities to develop evidence-based diagnostic criteria through computational analysis. This approach uses statistical analysis of large, structured datasets to identify the most diagnostically useful clinical features, translating subjective clinical pattern recognition into objective, quantifiable, and testable diagnostic standards.

This data-driven approach offers several advantages: standardization provides consistent diagnostic criteria across different practitioners and settings; objectivity replaces reliance on expert opinion with statistical evidence; accessibility makes complex diagnostic expertise available to all practitioners; efficiency enables consistent application of validated multi-criterion decision rules that clinicians cannot reliably execute from memory under time pressure; and educational clarity helps practitioners understand which clinical features matter most for accurate diagnosis. Statistically validated disease phenotype lists can be provided to motivate clinicians to consider critically and document support for their tentative diagnoses.

In this manuscript, the term computational phenotyping refers specifically to diagnostic feature sets derived empirically from large-scale clinical data using statistical resampling and feature-importance analysis. This usage is distinct from consensus-based criteria, rule-enumeration systems, or a priori diagnostic algorithms. The focus of this study is not to evaluate or compare existing diagnostic frameworks, but to demonstrate a generalizable, data-driven method for deriving reproducible diagnostic profiles directly from structured clinical records.

## Methods

### Comprehensive Data Collection System

We implemented a custom web-based structured note-taking system specifically designed for statistical analysis [5,6]. This system was previously validated for real-time diagnostic assistance on 10 common TMD and OFP diagnoses, using Bayesian inference algorithms, demonstrating superior performance compared to traditional machine learning approaches in orofacial pain diagnosis [11]. For the current study, we utilized the same structured data collection framework but applied Monte Carlo statistical analysis to retrospectively develop evidence-based diagnostic criteria.

Between July 2022 and August 2026 (49 months), we prospectively collected comprehensive data from 1,501 consecutive patients, creating a large, structured dataset of TMD/OFP cases. This systematic approach eliminated the ambiguity inherent in traditional free-text clinical notes while ensuring every data point was captured in a format suitable for statistical analysis. All diagnoses were coded using standard medical classification (ICD-10) and confirmed by board-certified orofacial pain specialists. Board certification in Orofacial Pain is conferred by the American Board of Orofacial Pain (ABOP) following written and oral examination; all diagnosing clinicians in this dataset hold this credential and practice within the USC Herman Ostrow School of Dentistry Orofacial Pain and TMD Clinic. The 126-category diagnostic space reflects standard ICD-10 classification supplemented by the DC/TMD (Diagnostic Criteria for Temporomandibular Disorders) taxonomy, not a study-specific schema.

The USC Division of Orofacial Pain”oper’tes a dual-mission clinic. TMD and orofacial pain evaluations and treatment comprise the primary clinical mission conducted across all clinic sessions. A separate, designated session is devoted to oral medicine and sleep medicine consultations, which constitute a diagnostically and operationally distinct patient population. For this study, cases were restricted to the TMD/OFP treatment population. Patients seen exclusively for oral medicine evaluation, oral pathology workup, infectious oral conditions, bone pathology unrelated to TMJ, or sleep disorder management were excluded from the analytical dataset. This exclusion is both scientifically appropriate—these conditions address different diagnostic questions and would confound feature importance calculations derived from TMD/OFP presentations—and operationally straightforward, as the clinic scheduling system distinguishes the two populations. After applying these exclusions, the analytical dataset comprised the 1,501 consecutive TMD/OFP patients who carried at least one final orofacial pain diagnosis.

### Selecting Conditions and Features for Analysis

From the original 126 diagnostic categories in our database, only conditions with at least 24 exemplar cases were included to ensure sufficient statistical power for reliable analysis. This yielded 15 conditions suitable for developing diagnostic profiles. Two further conditions met the case-count threshold but produced no usable profile and are reported as null results: cervical myalgia, in which no feature met the inclusion rule, and daytime clenching, which retained a single feature that was equally common in masticatory myalgia and sleep bruxism. Two codes for trismus (R25.2 and R25.9) were merged into a single entity of 62 cases after they proved indistinguishable on opening, onset and co-occurrence. For each condition, we identified clinical features that appeared in at least 50% of the exemplar patients with that specific diagnosis, following established principles for developing clinical diagnostic tools [7]. This approach ensures we focus on the most characteristic features of each condition.

### Clinical Feature Categories

The structured data collection is organized into 4 categories of clinical features including:

- Location features (labeled “Location:”) specify the anatomical regions within the head and neck area where patients identify their symptoms or problems. This spatial information provides crucial diagnostic context, particularly for conditions that affect specific anatomical structures or follow particular distribution patterns.
- History of Present Illness features (labeled “HPI-”) encompass systematically assessed elements evaluated for every patient, including symptom severity, temporal pattern (continuous versus intermittent), frequency of occurrence for intermittent symptoms, duration of individual episodes, anatomical distribution (unilateral versus bilateral), time since first onset of symptoms, symptom character (descriptive qualities), and perceived causation factors as reported by the patient. To facilitate statistical analysis, we have found it useful to quantify all time-related items (e.g. age, duration) internally as log_10_(days).
- Extra Questions (labeled “Q:”) are additional inquiries prompted by the complaint category, allowing for condition-specific symptom exploration. For example, patients presenting with headache disorders would be asked specific questions about headache characteristics, triggers, and associated symptoms, while those with joint problems would be questioned about mechanical symptoms and functional limitations.
- Examination features (labeled “Exam:”) represent objective clinical assessments performed by the clinician, including visual inspection, measurements, auscultation and palpation.

#### Monte Carlo Statistical Analysis Framework

We employed Monte Carlo simulation with bootstrap resampling to determine the relative importance of identified clinical features for each of the 15 diagnoses. This methodology was particularly well-suited for diagnostic conditions with overlapping symptoms because the iterative bootstrap sampling provided robust estimates of feature importance while maintaining clinical interpretability. For each Monte Carlo iteration, we generated stratified bootstrap samples containing 80% of available cases while maintaining original case-to-control ratios to preserve natural prevalence patterns observed in clinical practice. Each bootstrap sample included stratified samples from all diagnostic categories, weighted by observed comorbidity patterns to reflect the complex relationships among different TMD conditions [8,10]. Within this framework, phenotypes are defined operationally as reproducible constellations of clinical features that demonstrate stable, statistically significant discriminative value across repeated resampling of real-world patient data.

Within each Monte Carlo iteration, we calculated feature importance using three complementary statistical methods to ensure that importance rankings were not dependent on any single statistical assumption. Information gain measured entropy reduction when datasets were split by each feature, providing insight into the discriminative power of individual clinical findings. Chi-square analysis assessed independence between features and diagnoses. Mutual information quantified feature-diagnosis dependence while accounting for non-linear relationships that might be missed by simpler statistical approaches.

The final importance score was calculated on a 0-100 scale and was derived as a weighted combination of three key components. Prevalence difference between the target diagnosis and all other diagnoses received a 40% weight, reflecting the fundamental importance of feature specificity in diagnostic decision-making. Diagnostic performance metrics combining sensitivity and specificity also received a 40% weight, emphasizing the practical utility of features in clinical practice. Positive likelihood ratio, normalized and capped at 10, contributed the remaining 20% weight, providing additional insight into the diagnostic value of positive findings.

#### Statistical Significance and Validation

For each feature, we performed 1,000 permutation tests by randomly shuffling diagnostic labels while maintaining feature values, creating a null distribution against which to test the observed importance scores. Features were considered statistically significant if their observed importance score exceeded the 95^th^ percentile of the permuted distribution, corresponding to p<0.05. We applied Benjamini-Hochberg correction to control for multiple testing across all features, ensuring that our significance thresholds remained valid despite the large number of comparisons.

Each Monte Carlo iteration incorporated a 5-fold stratified cross-validation to prevent overfitting and ensure generalizability of our results. Feature importance was calculated on 80% of each bootstrap sample, with the remaining 20% used to validate discriminative power through area-under-the-curve (AUC), sensitivity, specificity, and balanced accuracy metrics. This approach provided robust estimates of how well our feature rankings would perform on new, unseen patient data.

### Implementation and Convergence

All simulations were implemented in Python 3.13 using established statistical libraries including scikit-learn (v1.6.1) for machine learning functions, NumPy (v2.2.4) for numerical computations, SciPy (v1.16.0) for statistical tests, and Pandas (v2.2.3) for data manipulation. We performed 10,000 Monte Carlo iterations for each diagnosis, with random seeds to ensure complete reproducibility of results. The Monte Carlo method successfully converged for all 15 diagnoses, with feature importance rankings showing stability indicated by a coefficient of variation less than 5% across iterations.

Final diagnostic profiles were developed by ranking features according to their median importance scores across all 10,000 Monte Carlo iterations for each diagnosis. We classified features into distinct categories based on both their importance scores and stability across iterations. Primary features were defined as those in the top 15^th^ percentile with significance in greater than 90% of iterations, representing the most reliable core diagnostic criteria. Secondary features included those in the 70^th^-85^th^ percentile range with significance in greater than 75% of iterations, providing additional diagnostic confidence while maintaining adequate reliability. Features below the 70^th^ percentile or with insufficient stability were excluded from final diagnostic profiles to maintain focus on the most clinically useful characteristics.

#### Quality Assurance and Clinical Validation

We implemented comprehensive validation procedures to ensure both statistical accuracy and clinical relevance of our findings. Random feature sets were tested to confirm chance-level performance, validating that our analytical methods could distinguish meaningful patterns from statistical noise. We verified that features previously established in the orofacial pain literature achieved appropriately high importance scores, confirming our methodology’s ability to recognize clinically relevant patterns. All 15 diagnostic profiles underwent systematic review by two board-certified orofacial pain specialists to verify clinical plausibility and consistency with established diagnostic guidelines, ensuring that our statistically-derived profiles aligned with expert clinical knowledge and current best practices in TMD diagnosis.

#### Diagnostic Alignment Procedure

Diagnostic alignment was performed by systematically comparing diagnostic entities and defining features contained in the clinical symptom–examination table with the Diagnostic Criteria for Temporomandibular Disorders (DC-TMD), Version 2/6/2020 [11]. Each diagnosis in the clinical table was evaluated against DC-TMD requirements using both history-based (DC-TMD Symptom Questionnaire items) and clinical examination-based (DC-TMD Examination Form items) criteria. Only diagnoses present in both documents were included in the comparison.

For each overlapping diagnosis, alignment was classified as *complete* when all essential DC-TMD criteria, specifically pain location, modification by jaw function, and required examination findings, were clearly represented in Table 1. Alignment was classified as *partial* when a diagnosis corresponded conceptually to a DC-TMD category but lacked sufficient documentation to distinguish specific DC-TMD subtypes (e.g., absence of reported spreading or referred pain in myofascial pain subcategories). Diagnoses outside the scope of DC-TMD (e.g., bruxism, neuropathic pain conditions, tinnitus, and burning mouth disorder) were excluded from analysis.

**Table 1:**
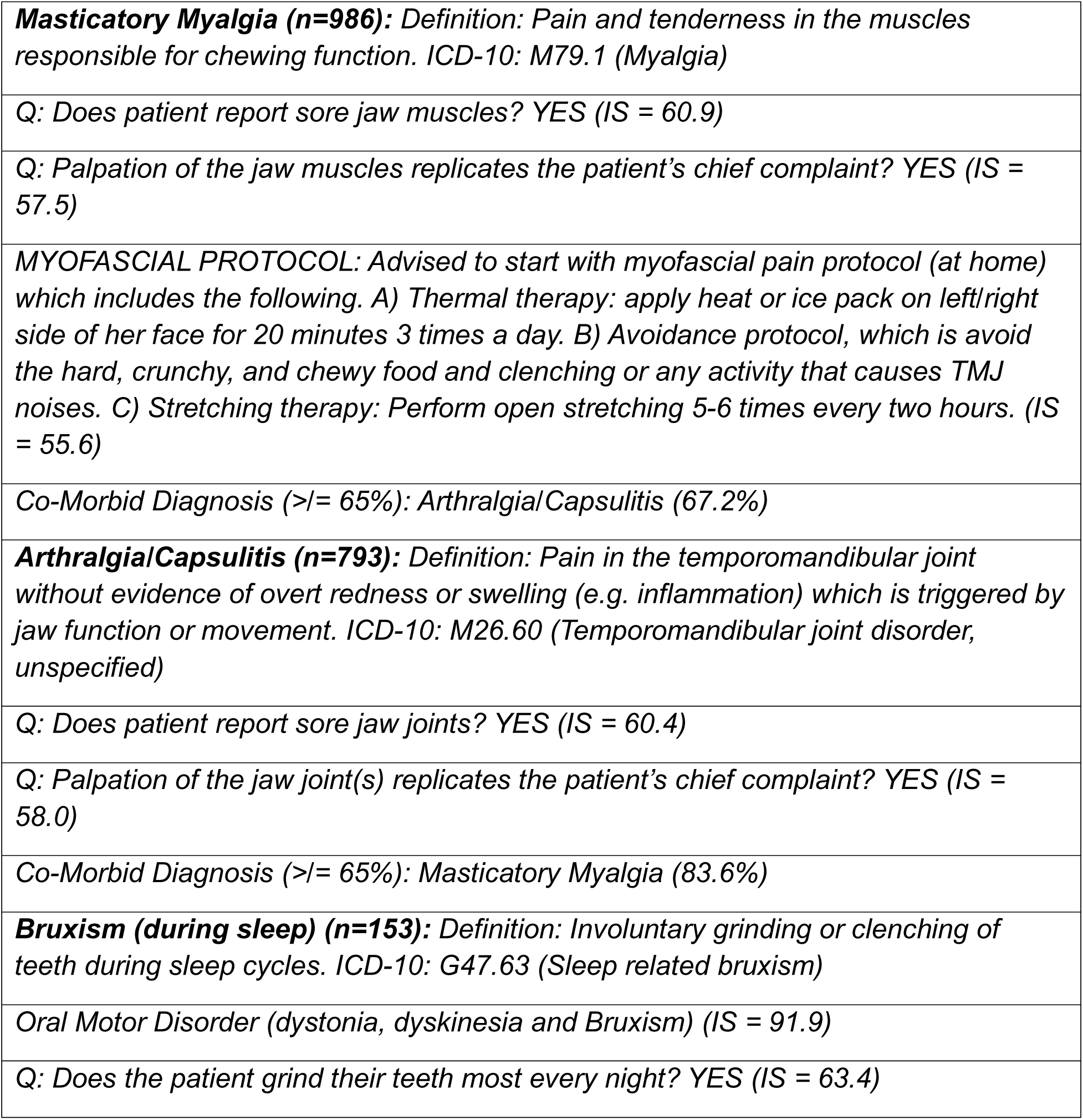
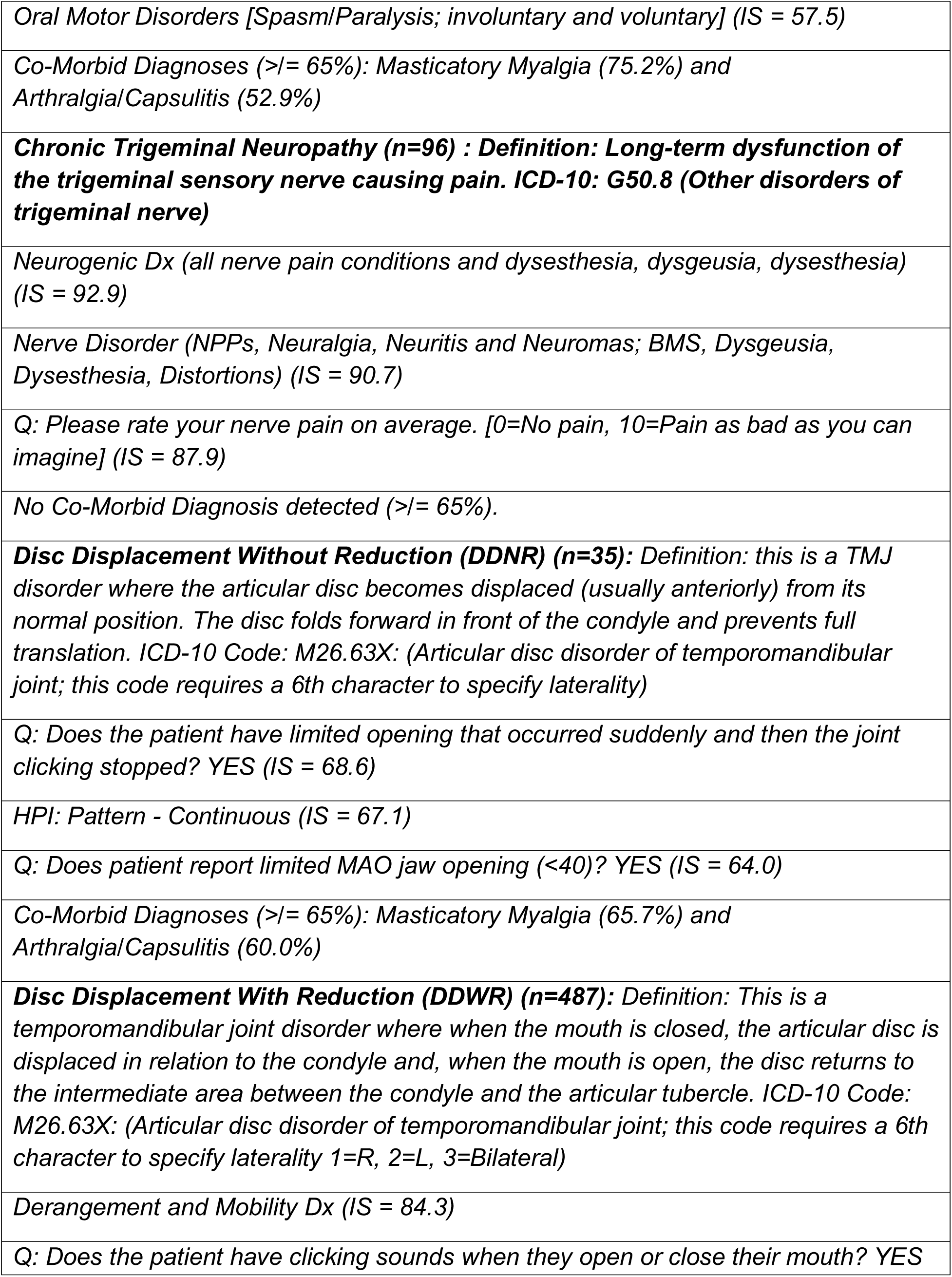
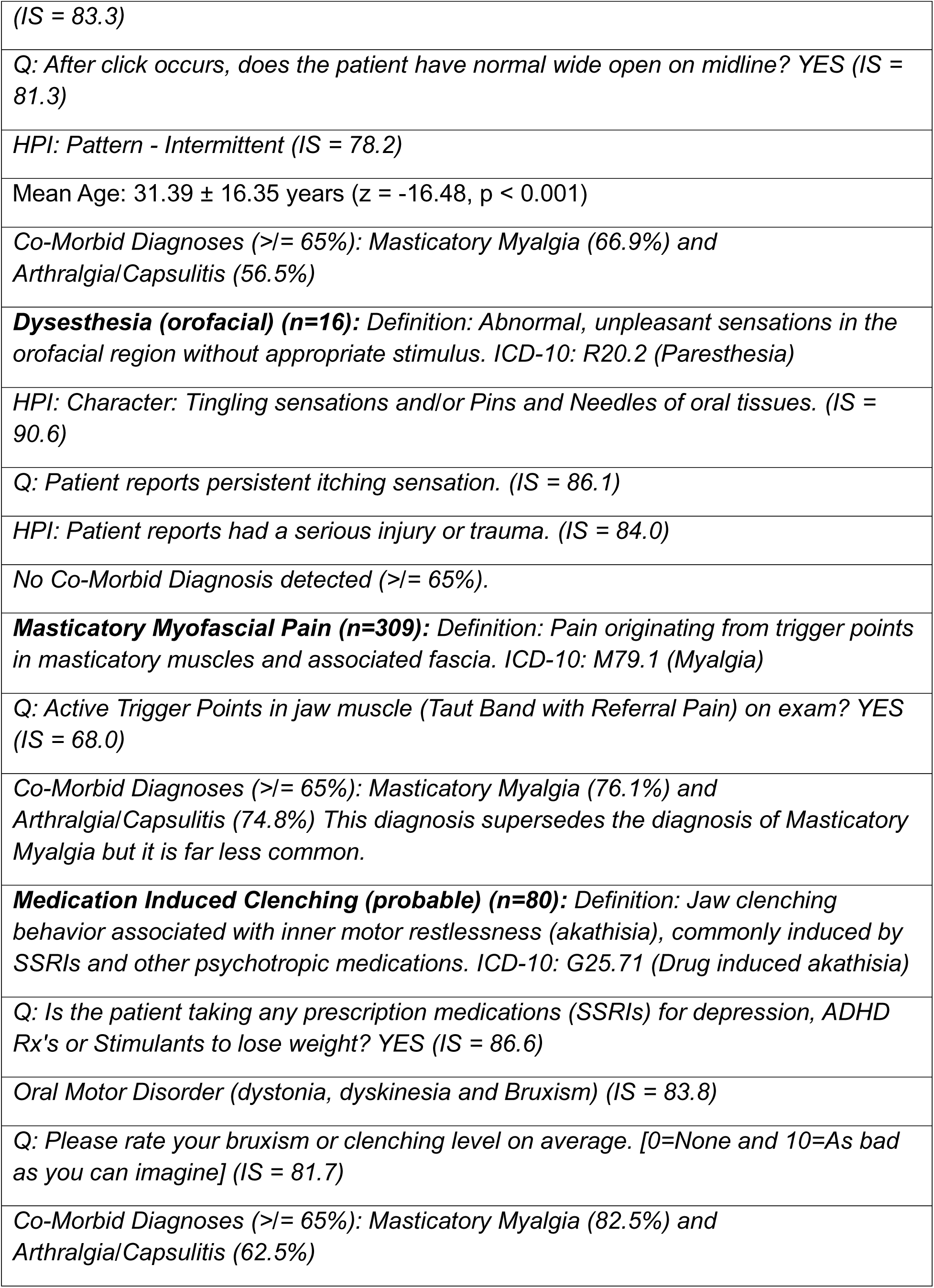
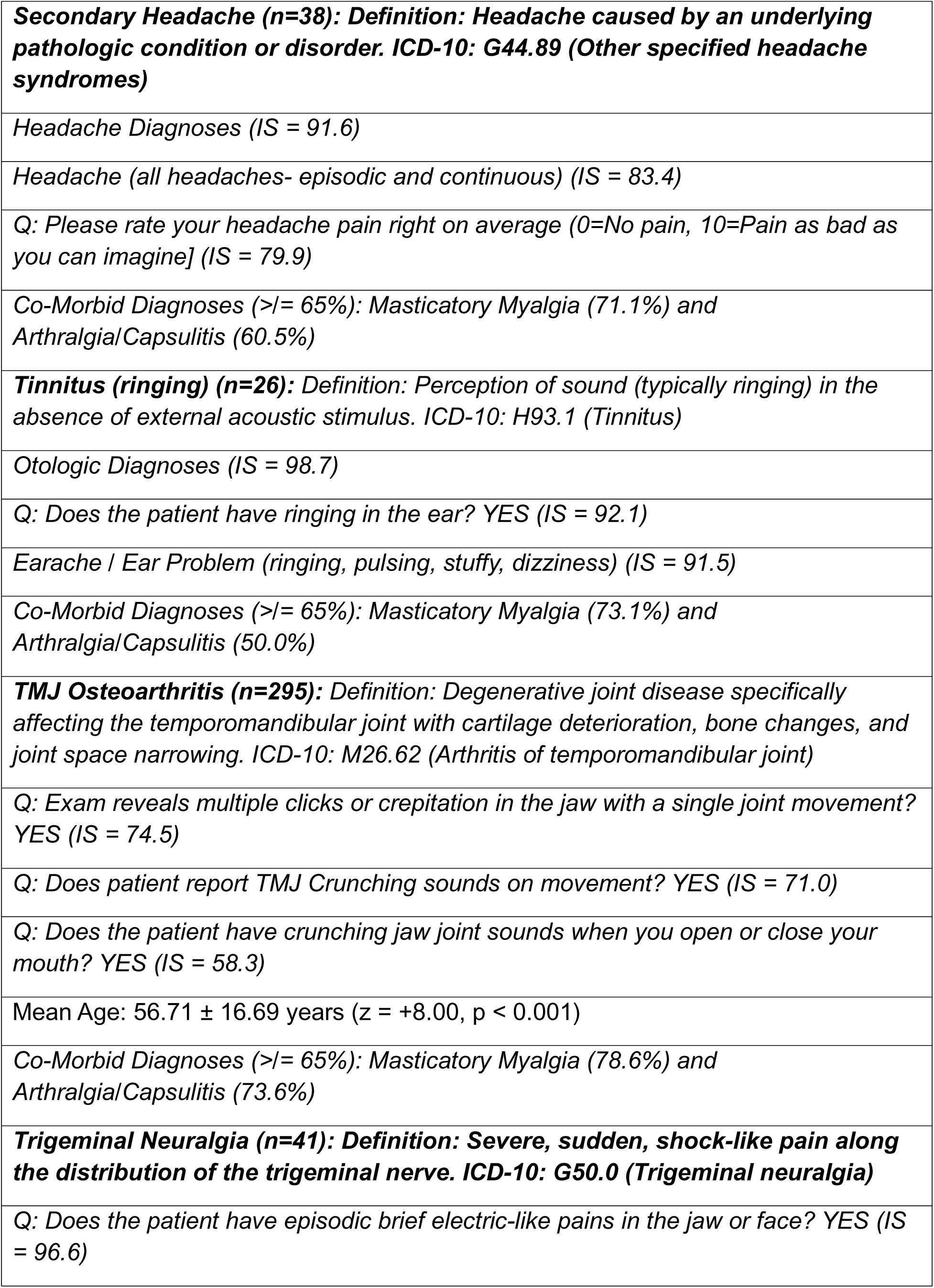
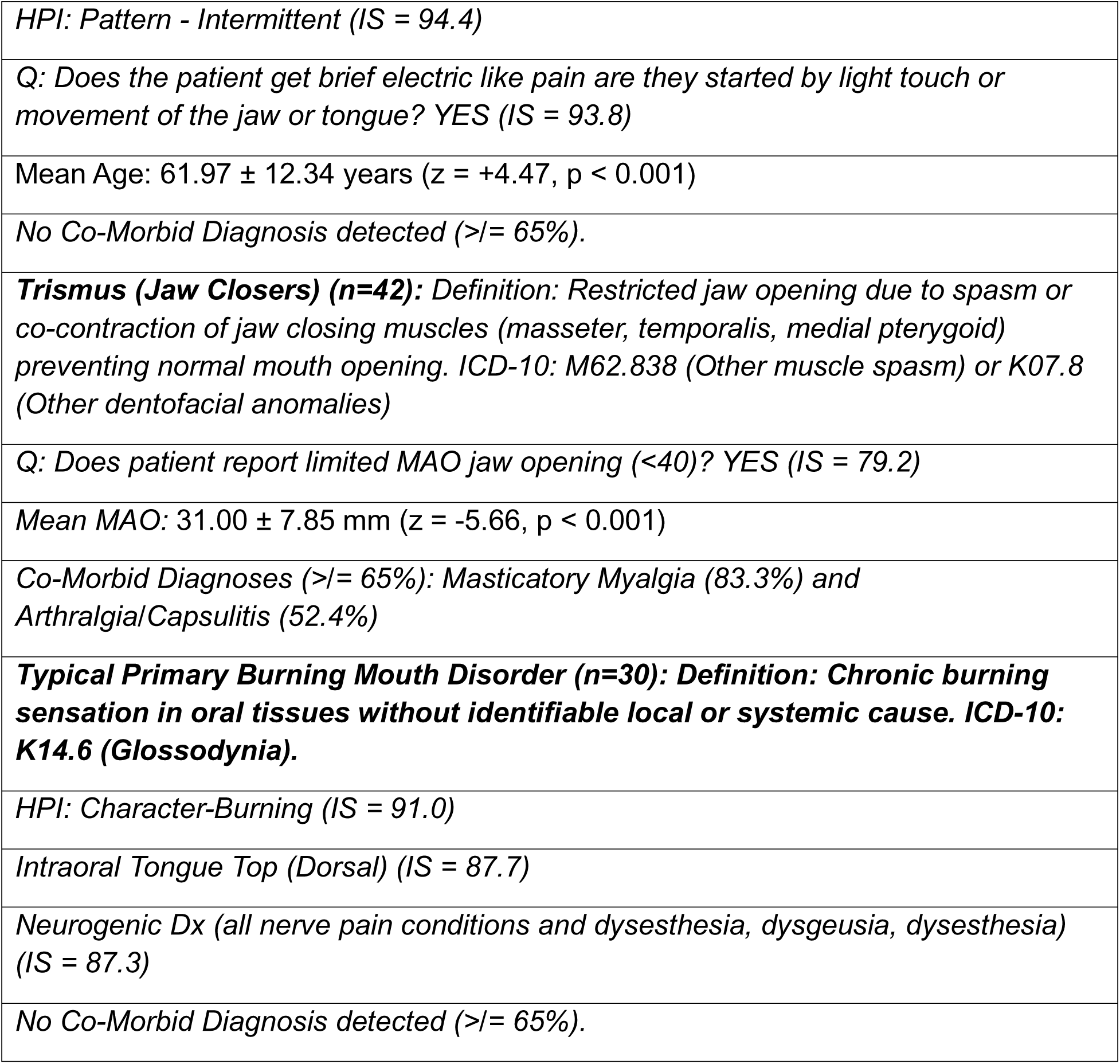
Computational Phenotypes with Monte Carlo Importance Scores. Below is a single table with an abbreviated list of feature lists for all 15 diagnoses. The full list of features for these diagnoses are available in Appendix 1. The features listed below are the top 3 to 8 features ranked by their Monte Carlo generated importance scores (IS). Where an individual diagnosis had a mean AGE or mean Maximum Active Opening (MAO) that was significantly different from the mean AGE or mean MAO of the entire dataset, it is also listed along with the z-score used in this analysis. In addition to these features, AGE and MAO, we list if one or more co-morbid diagnoses met our criteria (>/= 65%).

|  |
| --- |
| **Masticatory Myalgia (n=986):** Definition: Pain and tenderness in the muscles responsible for chewing function. ICD-10: M79.1 (Myalgia) |
| Q: Does patient report sore jaw muscles? YES (IS = 60.9) |
| Q: Palpation of the jaw muscles replicates the patient's chief complaint? YES (IS = 57.5) |
| MYOFASCIAL PROTOCOL: Advised to start with myofascial pain protocol (at home) which includes the following. A) Thermal therapy: apply heat or ice pack on left/right side of her face for 20 minutes 3 times a day. B) Avoidance protocol, which is avoid the hard, crunchy, and chewy food and clenching or any activity that causes TMJ noises. C) Stretching therapy: Perform open stretching 5-6 times every two hours. (IS = 55.6) |
| Co-Morbid Diagnosis ($\geq 65\%$): Arthralgia/Capsulitis (67.2%) |
| **Arthralgia/Capsulitis (n=793):** Definition: Pain in the temporomandibular joint without evidence of overt redness or swelling (e.g. inflammation) which is triggered by jaw function or movement. ICD-10: M26.60 (Temporomandibular joint disorder, unspecified) |
| Q: Does patient report sore jaw joints? YES (IS = 60.4) |
| Q: Palpation of the jaw joint(s) replicates the patient's chief complaint? YES (IS = 58.0) |
| Co-Morbid Diagnosis ($\geq 65\%$): Masticatory Myalgia (83.6%) |
| **Bruxism (during sleep) (n=153):** Definition: Involuntary grinding or clenching of teeth during sleep cycles. ICD-10: G47.63 (Sleep related bruxism) |
| Oral Motor Disorder (dystonia, dyskinesia and Bruxism) (IS = 91.9) |
| Q: Does the patient grind their teeth most every night? YES (IS = 63.4) |
| *Oral Motor Disorders [Spasm/Paralysis; involuntary and voluntary] (IS = 57.5)* |
| *Co-Morbid Diagnoses (>= 65%): Masticatory Myalgia (75.2%) and Arthralgia/Capsulitis (52.9%)* |
| **Chronic Trigeminal Neuropathy (n=96) : Definition: Long-term dysfunction of the trigeminal sensory nerve causing pain. ICD-10: G50.8 (Other disorders of trigeminal nerve)** |
| *Neurogenic Dx (all nerve pain conditions and dysesthesia, dysgeusia, dysesthesia) (IS = 92.9)* |
| *Nerve Disorder (NPPs, Neuralgia, Neuritis and Neuromas; BMS, Dysgeusia, Dysesthesia, Distortions) (IS = 90.7)* |
| *Q: Please rate your nerve pain on average. [0=No pain, 10=Pain as bad as you can imagine] (IS = 87.9)* |
| *No Co-Morbid Diagnosis detected (>= 65%).* |
| **Disc Displacement Without Reduction (DDNR) (n=35):** Definition: this is a TMJ disorder where the articular disc becomes displaced (usually anteriorly) from its normal position. The disc folds forward in front of the condyle and prevents full translation. ICD-10 Code: M26.63X: (Articular disc disorder of temporomandibular joint; this code requires a 6th character to specify laterality) |
| *Q: Does the patient have limited opening that occurred suddenly and then the joint clicking stopped? YES (IS = 68.6)* |
| *HPI: Pattern - Continuous (IS = 67.1)* |
| *Q: Does patient report limited MAO jaw opening (<40)? YES (IS = 64.0)* |
| *Co-Morbid Diagnoses (>= 65%): Masticatory Myalgia (65.7%) and Arthralgia/Capsulitis (60.0%)* |
| **Disc Displacement With Reduction (DDWR) (n=487):** Definition: This is a temporomandibular joint disorder where when the mouth is closed, the articular disc is displaced in relation to the condyle and, when the mouth is open, the disc returns to the intermediate area between the condyle and the articular tubercle. ICD-10 Code: M26.63X: (Articular disc disorder of temporomandibular joint; this code requires a 6th character to specify laterality 1=R, 2=L, 3=Bilateral) |
| *Derangement and Mobility Dx (IS = 84.3)* |
| *Q: Does the patient have clicking sounds when they open or close their mouth? YES* |
| *(IS = 83.3)* |
| *Q: After click occurs, does the patient have normal wide open on midline? YES (IS = 81.3)* |
| *HPI: Pattern - Intermittent (IS = 78.2)* |
| *Mean Age: 31.39 ± 16.35 years (z = -16.48, p < 0.001)* |
| *Co-Morbid Diagnoses (>= 65%): Masticatory Myalgia (66.9%) and Arthralgia/Capsulitis (56.5%)* |
| ***Dysesthesia (orofacial) (n=16):*** Definition: Abnormal, unpleasant sensations in the orofacial region without appropriate stimulus. ICD-10: R20.2 (Paresthesia) |
| *HPI: Character: Tingling sensations and/or Pins and Needles of oral tissues. (IS = 90.6)* |
| *Q: Patient reports persistent itching sensation. (IS = 86.1)* |
| *HPI: Patient reports had a serious injury or trauma. (IS = 84.0)* |
| *No Co-Morbid Diagnosis detected (>= 65%).* |
| ***Masticatory Myofascial Pain (n=309):*** Definition: Pain originating from trigger points in masticatory muscles and associated fascia. ICD-10: M79.1 (Myalgia) |
| *Q: Active Trigger Points in jaw muscle (Taut Band with Referral Pain) on exam? YES (IS = 68.0)* |
| *Co-Morbid Diagnoses (>= 65%): Masticatory Myalgia (76.1%) and Arthralgia/Capsulitis (74.8%) This diagnosis supersedes the diagnosis of Masticatory Myalgia but it is far less common.* |
| ***Medication Induced Clenching (probable) (n=80):*** Definition: Jaw clenching behavior associated with inner motor restlessness (akathisia), commonly induced by SSRIs and other psychotropic medications. ICD-10: G25.71 (Drug induced akathisia) |
| *Q: Is the patient taking any prescription medications (SSRIs) for depression, ADHD Rx's or Stimulants to lose weight? YES (IS = 86.6)* |
| *Oral Motor Disorder (dystonia, dyskinesia and Bruxism) (IS = 83.8)* |
| *Q: Please rate your bruxism or clenching level on average. [0=None and 10=As bad as you can imagine] (IS = 81.7)* |
| *Co-Morbid Diagnoses (>= 65%): Masticatory Myalgia (82.5%) and Arthralgia/Capsulitis (62.5%)* |
| **Secondary Headache (n=38): Definition: Headache caused by an underlying pathologic condition or disorder. ICD-10: G44.89 (Other specified headache syndromes)** |
| Headache Diagnoses (IS = 91.6) |
| Headache (all headaches- episodic and continuous) (IS = 83.4) |
| Q: Please rate your headache pain right on average (0=No pain, 10=Pain as bad as you can imagine] (IS = 79.9) |
| Co-Morbid Diagnoses (>= 65%): Masticatory Myalgia (71.1%) and Arthralgia/Capsulitis (60.5%) |
| **Tinnitus (ringing) (n=26): Definition: Perception of sound (typically ringing) in the absence of external acoustic stimulus. ICD-10: H93.1 (Tinnitus)** |
| Otologic Diagnoses (IS = 98.7) |
| Q: Does the patient have ringing in the ear? YES (IS = 92.1) |
| Earache / Ear Problem (ringing, pulsing, stuffy, dizziness) (IS = 91.5) |
| Co-Morbid Diagnoses (>= 65%): Masticatory Myalgia (73.1%) and Arthralgia/Capsulitis (50.0%) |
| **TMJ Osteoarthritis (n=295): Definition: Degenerative joint disease specifically affecting the temporomandibular joint with cartilage deterioration, bone changes, and joint space narrowing. ICD-10: M26.62 (Arthritis of temporomandibular joint)** |
| Q: Exam reveals multiple clicks or crepitation in the jaw with a single joint movement? YES (IS = 74.5) |
| Q: Does patient report TMJ Crunching sounds on movement? YES (IS = 71.0) |
| Q: Does the patient have crunching jaw joint sounds when you open or close your mouth? YES (IS = 58.3) |
| Mean Age: 56.71 ± 16.69 years (z = +8.00, p < 0.001) |
| Co-Morbid Diagnoses (>= 65%): Masticatory Myalgia (78.6%) and Arthralgia/Capsulitis (73.6%) |
| **Trigeminal Neuralgia (n=41): Definition: Severe, sudden, shock-like pain along the distribution of the trigeminal nerve. ICD-10: G50.0 (Trigeminal neuralgia)** |
| Q: Does the patient have episodic brief electric-like pains in the jaw or face? YES (IS = 96.6) |
| *HPI: Pattern - Intermittent (IS = 94.4)* |
| *Q: Does the patient get brief electric like pain are they started by light touch or movement of the jaw or tongue? YES (IS = 93.8)* |
| *Mean Age: 61.97 ± 12.34 years (z = +4.47, p < 0.001)* |
| *No Co-Morbid Diagnosis detected (>= 65%).* |
| ***Trismus (Jaw Closers) (n=42): Definition: Restricted jaw opening due to spasm or co-contraction of jaw closing muscles (masseter, temporalis, medial pterygoid) preventing normal mouth opening. ICD-10: M62.838 (Other muscle spasm) or K07.8 (Other dentofacial anomalies)*** |
| *Q: Does patient report limited MAO jaw opening (<40)? YES (IS = 79.2)* |
| *Mean MAO: 31.00 ± 7.85 mm (z = -5.66, p < 0.001)* |
| *Co-Morbid Diagnoses (>= 65%): Masticatory Myalgia (83.3%) and Arthralgia/Capsulitis (52.4%)* |
| ***Typical Primary Burning Mouth Disorder (n=30): Definition: Chronic burning sensation in oral tissues without identifiable local or systemic cause. ICD-10: K14.6 (Glossodynia).*** |
| *HPI: Character-Burning (IS = 91.0)* |
| *Intraoral Tongue Top (Dorsal) (IS = 87.7)* |
| *Neurogenic Dx (all nerve pain conditions and dysesthesia, dysgeusia, dysesthesia) (IS = 87.3)* |
| *No Co-Morbid Diagnosis detected (>= 65%).* |

## Results

### Dataset Characteristics

The analysis included 1,501 patients carrying 3,981 final diagnoses (mean 2.65 diagnoses per patient) drawn from 79 distinct diagnostic categories, with a mean age of 44.5 ± 20.4 years and a female representation of 75.2%. From the 126 diagnostic categories, 15 conditions met the inclusion criteria, with case volumes ranging from 16 to 986 cases per diagnosis.

### Diagnostic Performance Metrics

Cross-validation demonstrated discriminative performance across all 15 diagnostic categories, with AUC values ranging from 0.64 to 0.96. Feature selection was repeated within each training fold, so these are within-fold estimates. The highest performing conditions were Tinnitus (ringing) (AUC 0.96), Chronic Trigeminal Neuropathy (0.94), Medication Induced Clenching (0.94) and Trigeminal Neuralgia (0.92). The lowest performing conditions were TMJ Osteoarthritis (0.64) and Arthralgia/Capsulitis (0.66).

### Feature Importance and Stability

Of the 367 candidate features evaluated across the 15 diagnoses, 305 (83%) achieved statistical significance (p < 0.05) after Benjamini-Hochberg correction for multiple testing. Applying the inclusion rule left 109 features. All 109 retained their significance in more than 90% of the 10,000 bootstrap iterations, and the median coefficient of variation for their importance scores was 0.05, indicating robust convergence.

### Age and Functional Characteristics

Significant age differences from the database mean (p<0.05) were identified for several conditions. The youngest mean ages were observed in Disc Displacement With Reduction (31.6 ± 16.6 years) and Disc Displacement No Reduction (36.0 ± 17.1 years), against a cohort mean of 44.5 ± 20.4 years. The oldest mean ages occurred in Trigeminal Neuralgia (63.1 ± 10.9 years), TMJ Osteoarthrosis (59.1 ± 20.5 years), Chronic Trigeminal Neuropathy (58.4 ± 14.6 years) and TMJ Osteoarthritis (56.9 ± 17.1 years). Maximum active opening (cohort mean 44.2 ± 8.9 mm) was significantly reduced in Jaw Closer Trismus (30.0 ± 6.8 mm) and Disc Displacement No Reduction (34.0 ± 8.9 mm).

### Comorbidity Patterns

Complex comorbidity patterns above our arbitrary threshold of 65% emerged across multiple diagnostic categories. Masticatory Myalgia was the most frequently comorbid condition, reaching the 65% threshold in 10 of the other 14 diagnoses. Arthralgia/Capsulitis was the only other condition to reach the threshold, doing so in 3 of the other 14 diagnoses.

## Discussion

### From Statistical Phenotypes to Clinical Reality: The Role of Algorithmic Assistance

Computational phenotyping should not be interpreted as a replacement for clinical judgment or experiential expertise. Rather, it provides a transparent, statistically grounded representation of diagnostic reasoning that clinicians already employ intuitively. By quantifying feature importance and stability, this approach makes expert reasoning explicit, testable, and amenable to continuous refinement as additional clinical data accrue. While these computational phenotypes provide statistically robust diagnostic criteria, the extensive quantitative detail may initially appear overwhelming. Even experienced practitioners cannot realistically memorize and systematically apply importance scores across all diagnostic categories. Most clinicians probably have mental patterns for the 10 most common conditions, but certainly not for all 126 TMD/OFP diagnoses. This represents a fundamental human cognitive limitation rather than a methodological shortcoming. The real clinical value lies in integrating these profiles into algorithmic diagnostic assistance systems where computers can instantly access, weight, and combine statistically validated criteria during patient encounters. As clinical decision-support systems advance, evidence-based disease phenotypes such as these will serve as the foundational knowledge base that enables accurate, consistent diagnostic suggestions. Such technology system will democratize access to specialized diagnostic knowledge regardless of practitioner expertise.

### The Diagnostic Jigsaw: Why Individual Symptoms Are Insufficient

On a cautionary note, clinicians must resist diagnosing based on isolated clinical findings, akin to identifying a jigsaw puzzle picture from only a few of its pieces. Although burning pain receives one of the highest importance score (94.1) for Typical Primary Burning Mouth Disorder, reliable and compelling diagnosis requires the complete phenotypic profile including burning character (94.1) and specific dorsal tongue location (77.7). Similarly, while limited mouth opening characterizes Disc Displacement No Reduction (93.5), accurate diagnosis demands additional features including firm end feel, unilateral side (84.8), and positive TMJ palpation (78.5). The strength lies in the constellation of multiple statistically validated criteria that create a comprehensive diagnostic picture, helping clinicians avoid premature diagnostic closure.

### Methodological Foundation: Why Monte Carlo Analysis Was Essential

Monte Carlo simulation with bootstrap resampling was chosen over traditional Gini importance scores due to critical limitations making Gini unsuitable for diagnostic phenotyping with small, imbalanced samples. Gini importance suffers from instability with small sample sizes and provides no statistical significance testing. As demonstrated by Strobl et al., Gini scores show a systematic preference for high-cardinality features (features with many unique values), which can lead to misleading feature importance rankings, potentially overlooking clinically meaningful indicators [9]. As established by Peduzzi and colleagues, Monte Carlo methods offer particularly attractive solutions for clinical investigators dealing with complex problems and limited sample sizes that are not amenable to customary mathematical approaches [10], and the feature selection represents a robust and validated methodology for identifying the relative importance of phenotypic features when attempting disease subphenotyping [12]. When used properly, these methods ensure that our diagnostic profiles reflect real-world patient presentations rather than algorithmic artifacts.

### Transforming Clinical Practice Through Evidence-Based Criteria

This study advances objective, evidence-based diagnostic criteria for conditions historically relying on subjective assessment. The 15 computational phenotypes presented here provide a rigorous statistical basis for traditional expert opinion. The exceptional stability of primary diagnostic features across 10,000 iterations provides strong evidence that these represent genuine diagnostic profiles, giving clinicians confidence in their consistency across different patient populations. As our database of structured EMR records grows and with the continuing clinical use of SmartNote, the statistical methods described herein can hopefully be extended automatically to the remaining 111 TMD/OFP diagnoses.

The diagnostic alignment analysis (Table 2) demonstrates that our computational phenotypes achieve complete alignment with established DC-TMD criteria for seven major diagnostic categories, validating the clinical utility of our data-driven approach. This concordance suggests that statistically derived phenotypes can complement and support existing expert-consensus frameworks, offering an evidence-based pathway for refining and updating diagnostic standards as clinical databases expand.

**Table 2.**
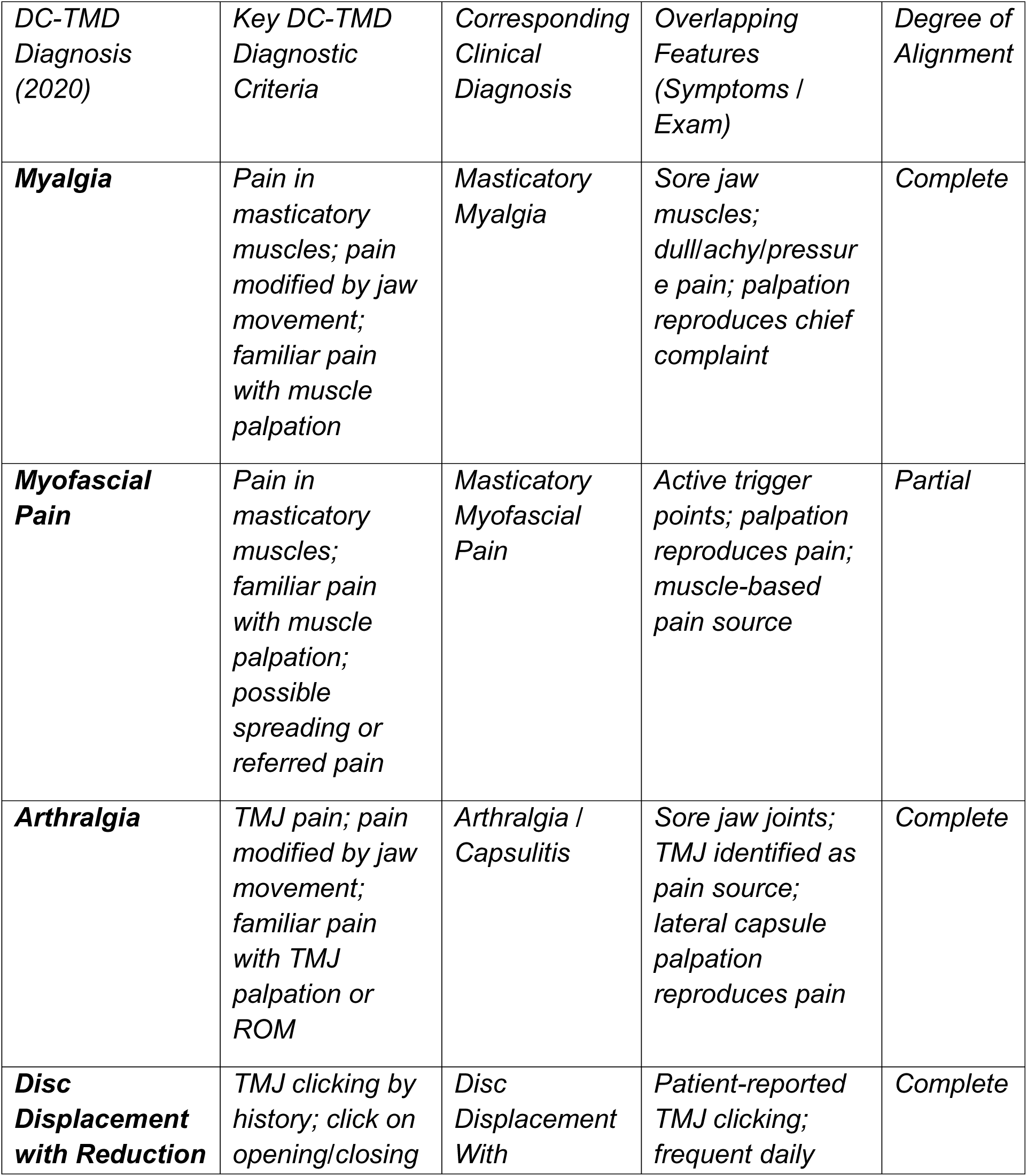
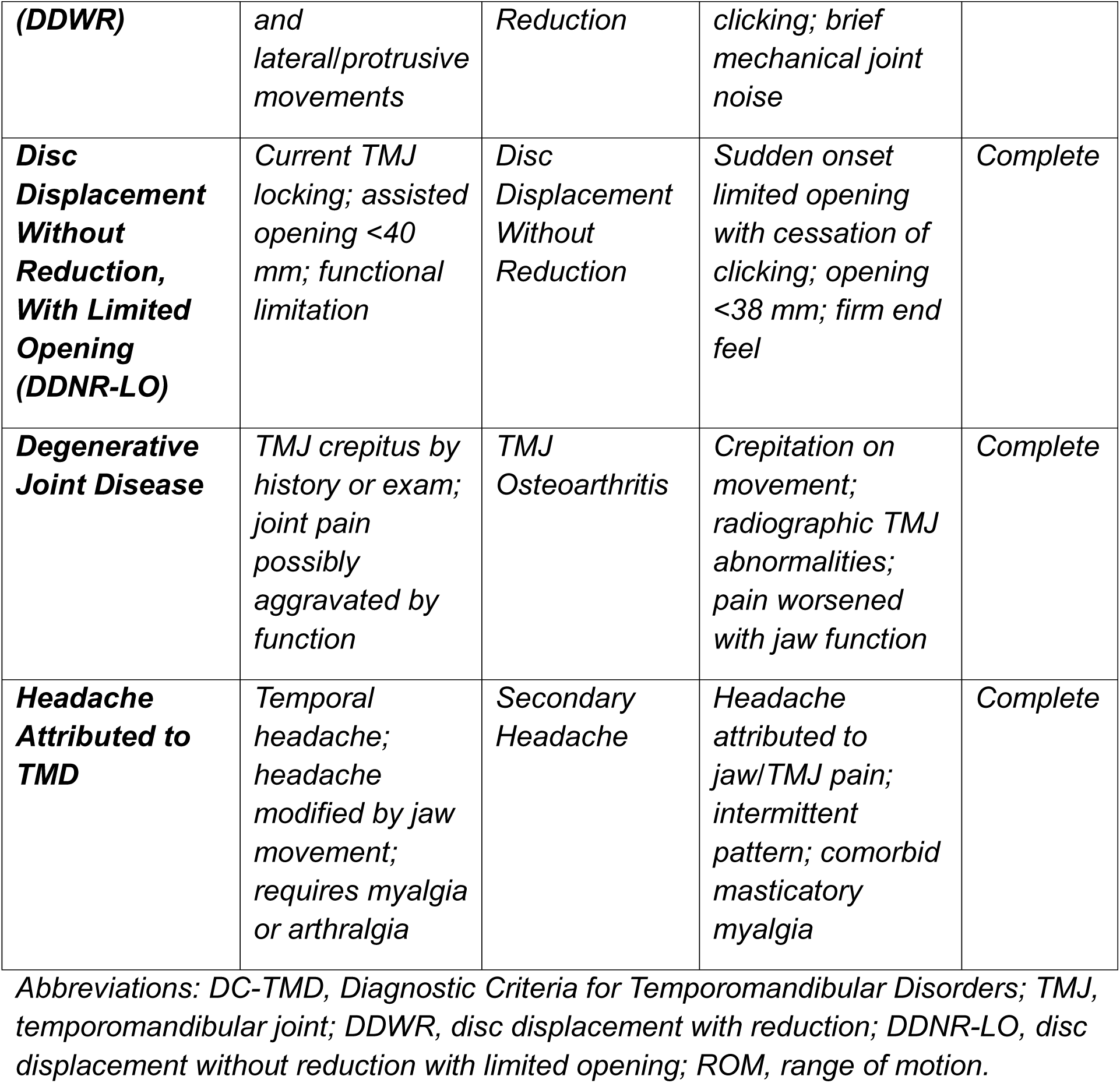
Alignment of DC-TMD (2020) Diagnostic Criteria with Corresponding Clinical Diagnoses. Compares diagnostic entities defined by the Diagnostic Criteria for Temporomandibular Disorders (DC-TMD), Version 2/6/2020, with corresponding diagnoses and clinical features reported in the attached symptom–examination table. Only diagnoses present in both Table 1 and DC-TMD are included. Alignment was determined based on overlap in required history items (DC-TMD Symptom Questionnaire) and clinical examination findings (DC-TMD Examination Form). “Complete” alignment indicates that the clinical table contains all essential DC-TMD diagnostic elements for that disorder. “Partial” alignment suggests that a diagnosis corresponds conceptually but lacks sufficient information to fully subtype the DC-TMD category (e.g., absence of documented spreading or referred pain for myofascial pain subtypes). Diagnoses present in Table 1 but outside the DC-TMD diagnostic framework (e.g., bruxism, neuropathic pain, tinnitus, burning mouth disorder) were intentionally excluded.

### Practical Applications

These profiles offer immediate benefits by codifying the mental checklists clinicians use intuitively to evaluate patients. The quantitative importance scores help practitioners understand which features carry the most diagnostic weight, supporting evidence-based education and skill development. Phenotype lists pop-ups can be presented to clinicians when they have identified a tentative diagnosis to encourage critical thinking and complete documentation of observations that support or question that diagnosis.

### Disease Interconnections and Clinical Insights

Interestingly, masticatory myalgia appearing as a comorbid condition in 11 of the other 14 diagnostic categories provides valuable insights into TMD/OFP interconnections, suggesting common underlying mechanisms including parafunction, central sensitization, and shared innervation pathways. Age-related patterns—younger patients with disc displacement conditions, older patients with neurological conditions—confirm epidemiological insights guiding age-appropriate differential diagnosis and may lead to previously unrecognized diagnostic criteria.

### Future Directions and Integration

The structured data collection approach represents evolution toward sophisticated electronic health records with built-in algorithmic assistance. Combined with our previous Bayesian inference work for real-time diagnostic assistance [13], these computational phenotypes create a comprehensive framework for improving diagnostic accuracy and EMR completeness across all clinical expertise levels.

### Study Limitations

Our dataset represents cases from a single specialized clinic, potentially limiting generalizability to general dental practice. The Monte Carlo approach relies on structured data collection quality and completeness. Some conditions had relatively small sample sizes despite meeting inclusion criteria, and structured data requirements may present implementation challenges for practices without similar systems. Statistical extraction of phenotypes as performed here provides an efficient way to update those phenotypes as more EMRs accrue in the database and as new diagnoses or observation elements start to appear in newer EMRs. The self-curating nature of the structured EMR generated by SmartNote system we employ greatly facilitates frequent enhancements of scope and quality of phenotype lists. Future work should focus on validation in general practice settings, developing simplified versions for practice management software integration, and expanding datasets to include more diverse patient populations. Because diagnostic labels were assigned by experienced orofacial pain specialists, the resulting phenotypes necessarily reflect current expert diagnostic practice; however, the statistical framework presented here allows such expert-derived labels to be evaluated, challenged, and refined over time rather than treated as fixed ground truth. The DC/TMD Axis II psychosocial battery was not included as a predictive feature because our structured data collection captures clinical presentation elements rather than patient-reported psychosocial indices; the relationship between psychosocial comorbidity and diagnostic phenotype is a distinct scientific question warranting dedicated investigation.

## Conclusions

In this study, we developed 15 evidence-based diagnostic profiles for TMD and orofacial pain conditions through Monte Carlo analysis of 1,501 patient cases, achieving discriminative performance (AUC 0.64 to 0.96). These computational phenotypes, accompanied by quantified importance scores, offer statistically derived, objective diagnostic criteria that can function as practical checklists in clinical settings. More importantly, they provide a transparent foundation for algorithmic diagnostic support systems, supplying an evidence-based rationale for feature weighting that is currently missing in most AI-driven tools. Diagnostic alignment analysis confirms complete concordance between our computational phenotypes and DC-TMD criteria for seven major diagnostic categories, demonstrating that data-driven approaches can effectively complement and validate expert-consensus frameworks.

The future integration of these quantified diagnostic features into algorithmic classification models will enable real-time, evidence-based diagnostic assistance. For example, when clinicians document a highly specific symptom such as “brief electric shock pain” (importance score 98.1 for Trigeminal Neuralgia), algorithms can appropriately weight this feature, substantially increasing diagnostic accuracy. This establishes a synergistic relationship in which documenting high-importance features strengthens algorithmic confidence, while the absence of key features helps rule out competing conditions.

This methodology provides a scalable framework for other complex medical conditions and represents a meaningful step toward reducing diagnostic delays and errors, ultimately enhancing patient care through the fusion of evidence-based medicine and artificial intelligence.

## Data Availability

All data produced in the present study are available upon reasonable request to the authors

## Statement of IRB

The retrospective review and analysis of the dataset has been approved under the USC UPIIRB ##UP-07-00416/ Research project title: Retrospective chart review and data analysis of various Orofacial disorders seen in the Orofacial Pain and Oral Medicine Clinic.

## Acknowledgments

This work was supported internally by the Orofacial Pain and Oral Medicine Center at the Herman Ostrow School of Dentistry of USC. The findings and conclusions expressed herein are those of the authors.

## Appendix 1

*Below are 15 diagnoses with tables listing all phenotypic features that met the inclusion rule: a median Monte Carlo importance score of at least 50 out of 100 together with a positive likelihood ratio of at least 3. The table lists the cross-validation [mean area under the curve (AUC)] for these features. The table lists the those diagnoses that were co-morbid at a substantial level [>/=65%]. The phenotypic features are ranked by median importance score based on a 10,000 Monte Carlo iteration analysis. Prevalence of each feature, sensitivity/specificity and likelihood ratios are also listed. The importance scores range from 0.0-100.0 but only those >/= 50 with a positive likelihood ratio >/= 3 are included. Age and Maximum Active Opening measurements were not ranked but are included in the table when they were found to be significantly different from the mean age/MAO of the entire dataset of* 1,501 cases.

Myalgia (n=986)

Cross-validation AUC: 0.73

Co-Morbid:

TMJ Arthralgia: 67.2%

|  | **Clinical Feature** | **Prevalence (%)** | **Non-Prevalence (%)** | **Sensitivity (%)** | **Specificity (%)** | **LR+** | **Importance Score** |
| --- | --- | --- | --- | --- | --- | --- | --- |
| 1 | Does patient report sore jaw muscles?<br>YES | 83.2 | 25.8 | 83.2 | 74.2 | 3.24 | 60.9 |
| 2 | Palpation of the jaw muscles replicates the patient's chief complaint | 63.2 | 15.0 | 63.2 | 85.0 | 4.28 | 57.5 |

|  | Clinical Feature | Prevalence (%) | Non-Prevalence (%) | Sensitivity (%) | Specificity (%) | LR+ | Importance Score |
| --- | --- | --- | --- | --- | --- | --- | --- |
|  | ? YES |  |  |  |  |  |  |
| 3 | MYOFASCIAL PROTOCOL:<br>Advised to start with myofascial pain protocol (at home) which includes the following.<br>A)<br>Thermal therapy: apply heat or ice pack on left/right side of her face for 20 minutes 3 times a day. B)<br>Avoidance protocol, which is avoid the hard,<br>crunchy, and chewy food and clenching or any activity that causes TMJ noises. C) Stretching therapy: Perform open stretching 5-6 times every two hours. | 63.8 | 17.1 | 63.8 | 82.9 | 3.78 | 55.6 |
| 4 | Location: Jaw Muscles | 55.6 | 14.2 | 55.6 | 85.8 | 3.98 | 52.8 |

TMJ Arthralgia (n=793)

Cross-validation AUC: 0.66

Co-Morbid:

Myalgia: 83.6%

|  | **Clinical Feature** | **Prevalence (%)** | **Non-Prevalence (%)** | **Sensitivity (%)** | **Specificity (%)** | **LR +** | **Importance Score** |
| --- | --- | --- | --- | --- | --- | --- | --- |
| 1 | Does patient report sore jaw joints? YES | 78.1 | 22.5 | 78.1 | 77.6 | 3.50 | 60.4 |
| 2 | Palpation of the jaw joint(s) replicates the patient's chief complaint? YES | 56.0 | 10.6 | 56.0 | 89.4 | 5.38 | 58.0 |

DDWR (n=487)

Cross-validation AUC: 0.88

Co-Morbid:

Myalgia: 66.9%

TMJ Arthralgia: 56.5%

|  | **Clinical Feature** | **Prevalence (%)** | **Non-Prevalence (%)** | **Sensitivity (%)** | **Specificity (%)** | **LR+** | **Importance Score** |
| --- | --- | --- | --- | --- | --- | --- | --- |
| 1 | Derangement and Mobility Dx | 98.6 | 14.4 | 98.6 | 85.6 | 6.89 | 84.3 |
| 2 | Does the patient have clicking sounds when they open or close their mouth? YES | 80.9 | 7.8 | 80.9 | 92.2 | 9.71 | 83.3 |
| 3 | After click occurs, does the patient have normal wide open on midline? YES | 71.3 | 2.4 | 71.3 | 97.6 | 10.00 | 81.3 |
| 4 | Pattern - Intermittent | 73.3 | 7.6 | 73.3 | 92.4 | 9.40 | 78.2 |
| 5 | Frequency - One or more times a day | 62.2 | 5.4 | 62.2 | 94.6 | 9.89 | 73.9 |
| 6 | Duration - Seconds/Minute es | 61.2 | 5.4 | 61.2 | 94.6 | 9.86 | 73.2 |
| 7 | TMJ Dysfunction or Mobility Disorder (Joint noises, restrictions, hypermobility and locking problems including ankylosis) | 86.4 | 19.5 | 86.5 | 80.5 | 4.45 | 69.1 |
| 8 | Please rate your pain by selecting a number that best describes your pain ON AVERAGE." (0 = No pain → 10 = Pain as bad as you can imagine) | 81.5 | 19.2 | 81.5 | 80.8 | 4.26 | 65.9 |
| 9 | Cause - Unknown | 59.5 | 12.9 | 59.6 | 87.1 | 4.65 | 57.3 |

TMJ Arthritis (n=295)

Cross-validation AUC: 0.64

Co-Morbid:

Myalgia: 78.6%

TMJ Arthralgia: 73.6%

|  | **Clinical Feature** | **Prevalence (%)** | **Non-Prevalence (%)** | **Sensitivity (%)** | **Specificity (%)** | **LR+** | **Importance Score** |
| --- | --- | --- | --- | --- | --- | --- | --- |
| 1 | Exam reveals multiple clicks or crepitation in the jaw with a single joint movement ? YES | 61.7 | 4.1 | 61.7 | 95.9 | 10.00 | 74.5 |
| 2 | Does patient report TMJ Crunching sounds on movement ? YES | 55.9 | 4.2 | 55.9 | 95.8 | 9.98 | 71.0 |
| 3 | Does the patient have crunching jaw joint sounds when you open or | 35.3 | 3.3 | 35.2 | 96.7 | 9.57 | 58.3 |

|  | Clinical Feature | Prevalence (%) | Non-Prevalence (%) | Sensitivity (%) | Specificity (%) | LR+ | Importance Score |
| --- | --- | --- | --- | --- | --- | --- | --- |
|  | close your mouth?<br>YES |  |  |  |  |  |  |

Chronic Trigeminal Neuropathy (n=96)

Cross-validation AUC: 0.94

Co-Morbid:

|  | **Clinical Feature** | **Prevalence (%)** | **Non-Prevalence (%)** | **Sensitivity (%)** | **Specificity (%)** | **LR+** | **Importance Score** |
| --- | --- | --- | --- | --- | --- | --- | --- |
| 1 | Neurogenic Dx (all nerve pain conditions and dysesthesia, dysgeusia, dysesthesia) | 97.9 | 9.3 | 97.9 | 90.8 | 9.86 | 92.9 |
| 2 | Nerve Disorder (NPPs, Neuralgia, Neuritis and Neuromas ; BMS, Dysgeusia, Dysesthesia, Distortions) | 95.8 | 9.8 | 95.8 | 90.2 | 9.54 | 90.7 |
| 3 | Please rate your nerve pain on average. [0=No pain, 10=Pain as bad as you can imagine] | 91.7 | 9.7 | 91.7 | 90.3 | 9.36 | 87.9 |
| 4 | Side - Unilaterally | 85.4 | 7.3 | 85.5 | 92.8 | 9.97 | 86.9 |
| 5 | Pattern - Continuous | 83.3 | 5.8 | 83.3 | 94.2 | 10.00 | 86.5 |
| 6 | Does the patient have unilateral dentoalveolar pain? YES | 71.9 | 1.2 | 72.0 | 98.8 | 10.00 | 82.4 |
| 7 | Has the focal dental alveolar pain (intermittent or continuous) been persistent (lasts three months or longer)? YES | 67.7 | 0.9 | 67.8 | 99.2 | 10.00 | 80.1 |
| 8 | Cause - Unknown | 69.8 | 7.8 | 69.8 | 92.2 | 8.87 | 74.9 |
| 9 | Character - Dull / Achy / Pressure | 54.2 | 1.4 | 54.2 | 98.7 | 10.00 | 71.7 |
| 10 | By patient report is the area of focal dental alveolar pain sensitive to light touch (needs confirmation on exam)? YES | 51.0 | 0.5 | 51.1 | 99.5 | 10.00 | 70.4 |

MEDICATION INDUCED CLENCHING (n=80)

Cross-validation AUC: 0.94

Co-Morbid:

Myalgia: 82.5%

TMJ Arthralgia: 62.5%

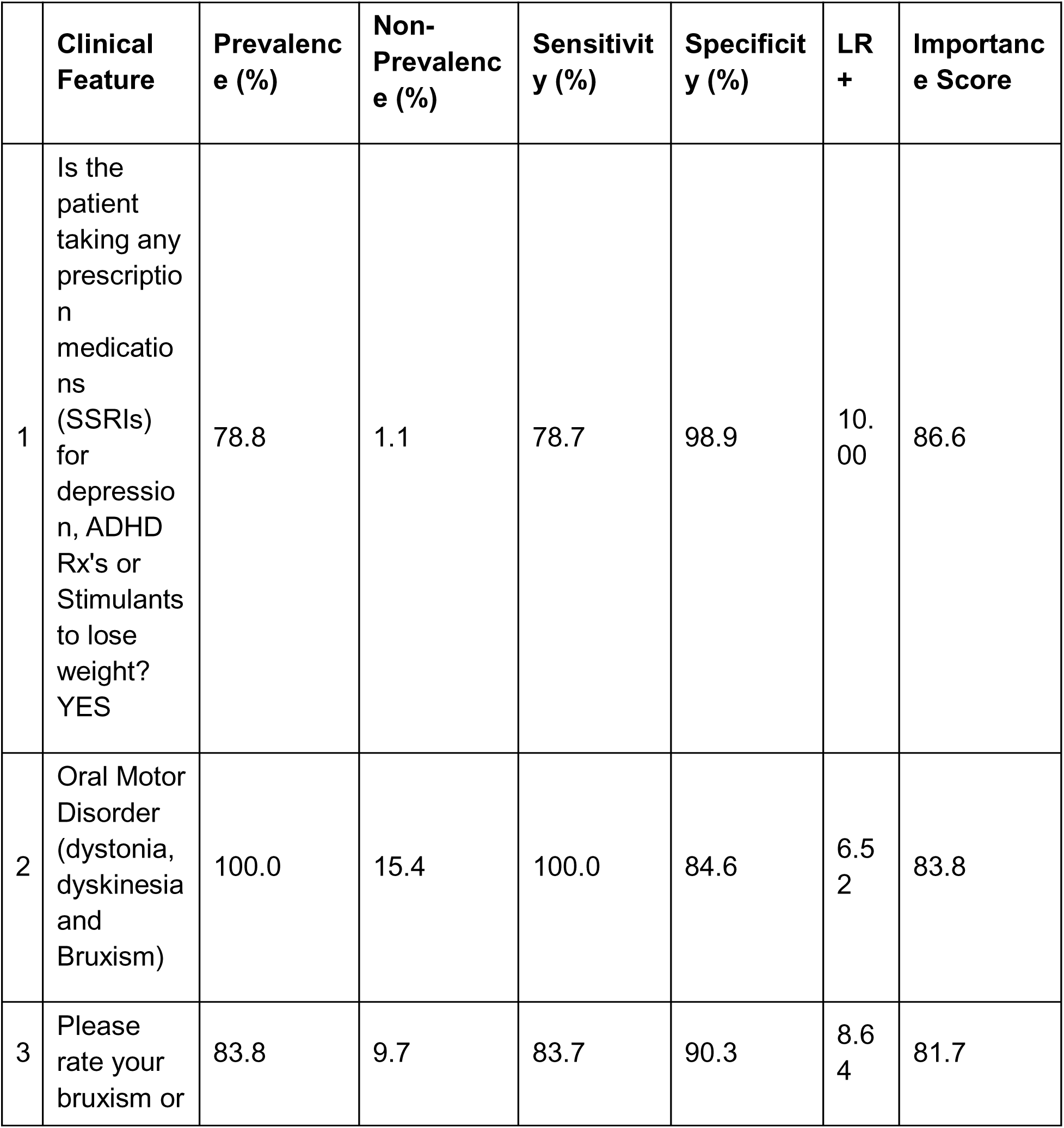
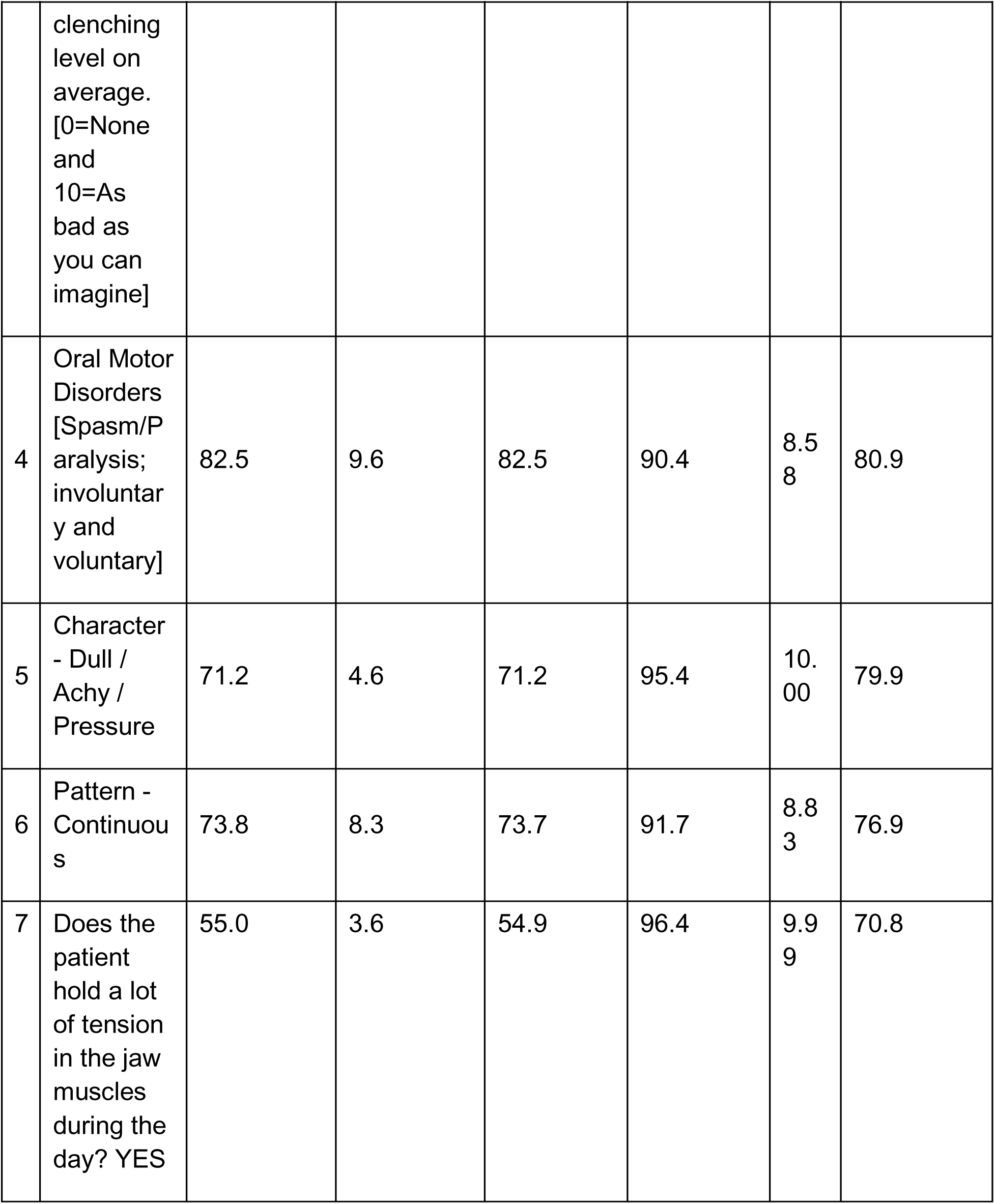
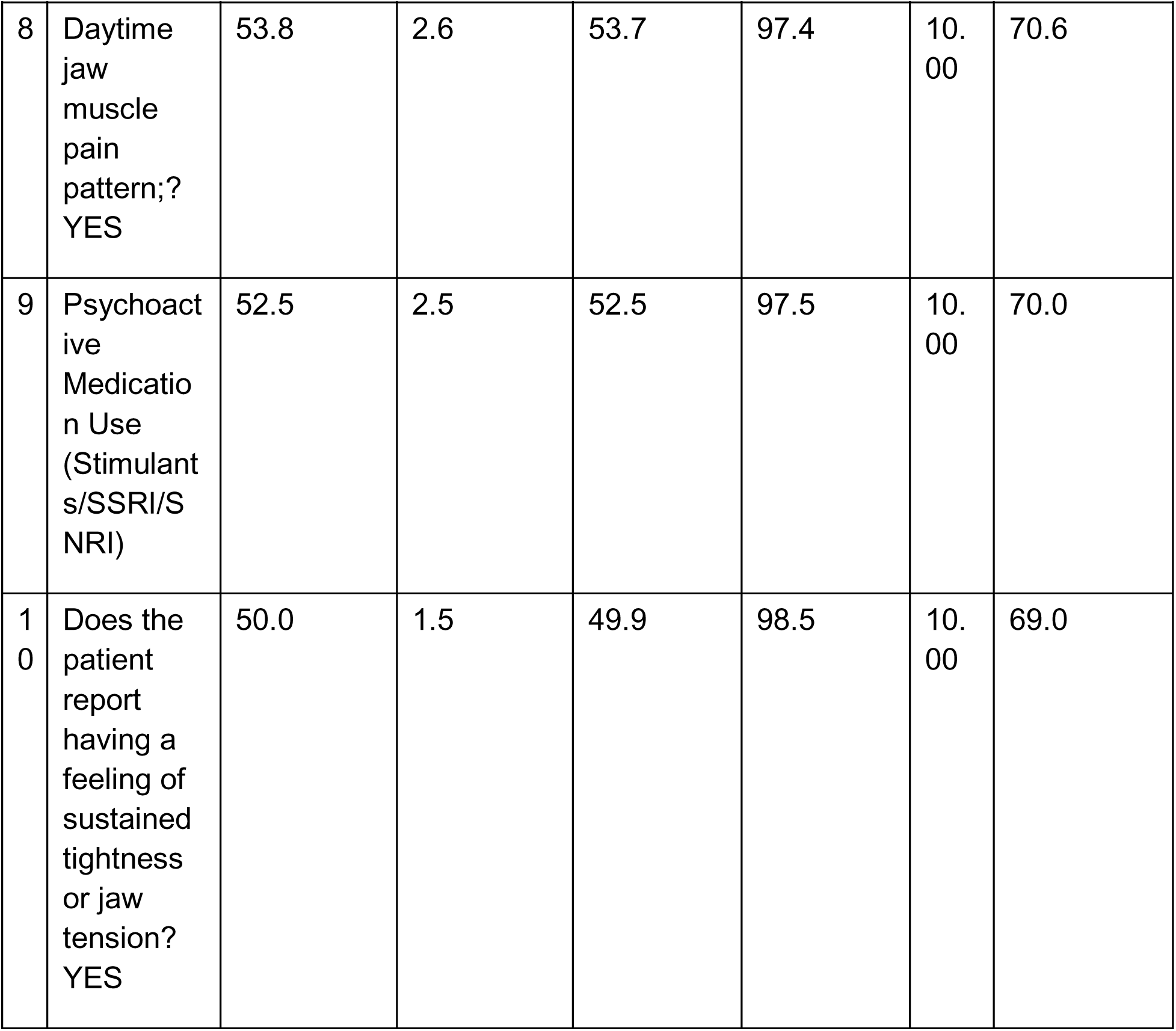

BRUXISM (n=153)

Cross-validation AUC: 0.9

Co-Morbid:

Myalgia: 75.2%

TMJ Arthralgia: 52.9%

|  | **Clinical Feature** | **Prevalence (%)** | **Non-Prevalence (%)** | **Sensitivity (%)** | **Specificity (%)** | **LR+** | **Importance Score** |
| --- | --- | --- | --- | --- | --- | --- | --- |
| 1 | Oral Motor Disorder (dystonia, dyskinesia and Bruxism) | 100.0 | 10.8 | 100.0 | 89.2 | 9.20 | 91.9 |
| 2 | Does the patient grind their teeth most every night?<br>YES | 41.2 | 2.2 | 41.2 | 97.8 | 10.00 | 63.4 |
| 3 | Oral Motor Disorders [Spasm/Paralysis; involuntary and voluntary] | 52.3 | 9.1 | 52.3 | 90.9 | 5.79 | 57.5 |
| 4 | Please rate your bruxism or clenching level on average. [0=None and 10=As bad as you can imagine] | 51.0 | 9.4 | 51.0 | 90.6 | 5.46 | 55.9 |
| 5 | Character - No pain | 27.5 | 1.8 | 27.5 | 98.2 | 9.95 | 55.3 |
| 6 | Side - Bilaterally | 39.9 | 5.6 | 39.9 | 94.4 | 7.26 | 55.1 |
| 7 | Cause - Clenching/Grinding | 28.8 | 2.6 | 28.7 | 97.4 | 9.56 | 54.8 |
| 8 | Onset - Years Ago | 38.6 | 5.3 | 38.6 | 94.7 | 7.41 | 54.8 |
| 9 | Patient has advanced attrition (shiny facets on Enamel surfaces)? YES | 53.6 | 14.2 | 53.6 | 85.8 | 3.81 | 51.3 |
| 10 | Does the patient have waking jaw pain? YES | 19.6 | 1.0 | 19.6 | 99.0 | 9.98 | 51.2 |

Trigeminal Neuralgia (n=41)

Cross-validation AUC: 0.92

Co-Morbid:

|  | **Clinical Feature** | **Prevalence (%)** | **Non-Prevalence (%)** | **Sensitivity (%)** | **Specificity (%)** | **LR +** | **Importance Score** |
| --- | --- | --- | --- | --- | --- | --- | --- |
| 1 | Does the patient have episodic brief electric-like pains in the jaw or face?<br>YES | 95.1 | 0.9 | 95.2 | 99.1 | 10.00 | 96.6 |
| 2 | Pattern - Intermittent | 92.7 | 2.1 | 92.7 | 97.9 | 10.00 | 94.4 |
| 3 | Does the patient get brief electric like pain are they started by light touch or movement of the jaw or tongue?<br>YES | 90.2 | 0.5 | 90.2 | 99.4 | 10.00 | 93.8 |
| 4 | Character - Electric | 90.2 | 0.7 | 90.2 | 99.3 | 10.00 | 93.7 |
| 5 | Duration - Seconds/Minutes | 87.8 | 1.1 | 87.8 | 98.9 | 10.00 | 92.0 |
| 6 | Side - Unilaterally | 95.1 | 9.9 | 95.1 | 90.1 | 9.42 | 90.0 |
| 7 | Neurogenic Dx (all nerve pain conditions and dysesthesia, dysgeusia, dysesthesia) | 100.0 | 12.5 | 100.0 | 87.5 | 8.03 | 88.5 |
| 8 | Nerve Disorder (NPPs, Neuralgia, Neuritis and Neuromas; BMS, Dysgeusia, Dysesthesia , Distortions) | 100.0 | 12.9 | 100.0 | 87.1 | 7.78 | 87.8 |
| 9 | Cause - Unknown | 90.2 | 9.6 | 90.2 | 90.4 | 9.28 | 86.9 |
| 10 | Does the patient have brief (<20 seconds) pain attacks?<br>YES | 78.0 | 0.2 | 78.0 | 99.8 | 10.00 | 86.7 |

TRIGEMINAL SENSORY DISORDER - DYSESTHESIA (n=16)

Cross-validation AUC: 0.95

Co-Morbid:

|  | **Clinical Feature** | **Prevalence (%)** | **Non-Prevalence (%)** | **Sensitivity (%)** | **Specificity (%)** | **LR +** | **Importance Score** |
| --- | --- | --- | --- | --- | --- | --- | --- |
| 1 | Character - Tingling / Pins and needles / Numbness | 85.7 | 1.4 | 85.8 | 98.6 | 10.00 | 90.6 |
| 2 | Pattern - Continuous | 91.4 | 7.7 | 91.4 | 92.3 | 9.97 | 90.1 |
| 3 | Side - Unilaterally | 88.6 | 8.3 | 88.6 | 91.8 | 9.84 | 87.9 |
| 4 | Does the patient have persistent itching sensation... | 77.1 | 0.2 | 77.0 | 99.8 | 10.00 | 86.1 |
| 5 | Cause - Trauma | 77.1 | 1.2 | 77.1 | 98.8 | 10.00 | 85.5 |
| 6 | Does the patient have persistent pain, tingling | 77.1 | 1.7 | 77.2 | 98.3 | 10.00 | 85.3 |
| 7 | Onset - Months Ago | 82.9 | 7.7 | 82.8 | 92.3 | 9.79 | 84.7 |
| 8 | Has the patient had a serious injury to trauma | 74.3 | 0.9 | 74.3 | 99.1 | 10.00 | 84.0 |
| 9 | Does the patient have a very uncomfortable bite | 74.3 | 2.9 | 74.3 | 97.1 | 10.00 | 82.8 |

Secondary Headache (n=38)

Cross-validation AUC: 0.9

Co-Morbid:

Myalgia: 71.1%

TMJ Arthralgia: 60.5%

|  | **Clinical Feature** | **Prevalence (%)** | **Non-Prevalence (%)** | **Sensitivity (%)** | **Specificity (%)** | **LR+** | **Importance Score** |
| --- | --- | --- | --- | --- | --- | --- | --- |
| 1 | Headache Diagnoses | 92.1 | 6.1 | 92.1 | 93.9 | 10.00 | 91.6 |
| 2 | Headache (all headache-episodic and continuous) | 92.1 | 12.0 | 92.1 | 88.0 | 7.70 | 83.4 |
| 3 | Please rate your headache pain right on average (0=No pain, 10=Pain as bad as you can imagine] | 86.8 | 11.8 | 86.8 | 88.2 | 7.43 | 79.9 |
| 4 | Side - Bilaterally | 73.7 | 8.2 | 73.7 | 91.8 | 8.86 | 77.0 |
| 5 | Character - Dull / Achy / Pressure | 73.7 | 10.1 | 73.6 | 89.9 | 7.33 | 72.8 |
| 6 | Does the headache involve pain and tightness in the temple and/or neck? YES | 63.2 | 7.0 | 63.2 | 93.0 | 8.84 | 71.4 |
| 7 | Onset - Years Ago | 60.5 | 6.2 | 60.5 | 93.8 | 9.23 | 71.1 |
| 8 | Duration - Hours | 57.9 | 6.6 | 57.9 | 93.4 | 8.62 | 68.0 |
| 9 | Pattern - Intermittent | 60.5 | 8.2 | 60.5 | 91.8 | 7.42 | 66.2 |

|  | Clinical Feature | Prevalence (%) | Non-Prevalence (%) | Sensitivity (%) | Specificity (%) | LR+ | Importance Score |
| --- | --- | --- | --- | --- | --- | --- | --- |
| 10 | Does the patient feel the headaches are being caused by your jaw or tooth or neck pain?<br>YES | 44.7 | 3.9 | 44.8 | 96.1 | 9.55 | 63.6 |

BMS (n=30)

Cross-validation AUC: 0.91

Co-Morbid:

|  | **Clinical Feature** | **Prevalence (%)** | **Non-Prevalence (%)** | **Sensitivity (%)** | **Specificity (%)** | **LR+** | **Importance Score** |
| --- | --- | --- | --- | --- | --- | --- | --- |
| 1 | Character-Burning | 86.7 | 1.7 | 86.8 | 98.3 | 10.00 | 91.0 |
| 2 | Intraoral Tongue Top (Dorsal) | 80.0 | 0.5 | 80.0 | 99.5 | 10.00 | 87.7 |
| 3 | Neurogenic Dx (all nerve pain conditions and dysesthesia, dysgeusia, dysesthesia) | 100.0 | 13.2 | 100.0 | 86.8 | 7.61 | 87.3 |
| 4 | Side - Bilaterally | 70.0 | 1.4 | 70.1 | 98.6 | 10.00 | 81.2 |
| 5 | Pattern - Continuous | 80.0 | 9.4 | 80.1 | 90.6 | 8.52 | 79.5 |
| 6 | Does the patient have a persistent burning sensation on the top side of the tongue?<br>YES | 63.3 | 0.1 | 63.5 | 99.9 | 10.00 | 78.0 |
| 7 | Please rate your nerve pain on average. [0=No pain, 10=Pain as bad as you can imagine] | 86.7 | 13.5 | 86.8 | 86.5 | 6.47 | 76.9 |
| 8 | Nerve Disorder (NPPs, Neuralgia, Neuritis and Neuromas; | 86.7 | 13.9 | 86.8 | 86.1 | 6.28 | 76.3 |

|  | Clinical Feature | Prevalence (%) | Non-Prevalence (%) | Sensitivity (%) | Specificity (%) | LR+ | Importance Score |
| --- | --- | --- | --- | --- | --- | --- | --- |
|  | BMS, Dysgeusia, Dysesthesia, Distortions) |  |  |  |  |  |  |
| 9 | Cause - Unknown | 73.3 | 10.5 | 73.4 | 89.5 | 7.01 | 71.8 |
| 10 | Intraoral Exam Findings | 83.3 | 17.5 | 83.3 | 82.5 | 4.78 | 69.1 |

CLOSER TRISMUS (n=42)

Cross-validation AUC: 0.77

Co-Morbid:

Myalgia: 83.3%

TMJ Arthralgia: 52.4%

|  | **Clinical Feature** | **Prevalence (%)** | **Non-Prevalence (%)** | **Sensitivity (%)** | **Specificity (%)** | **LR+** | **Importance Score** |
| --- | --- | --- | --- | --- | --- | --- | --- |
| 1 | Does patient report limited MAO jaw opening (<40)? YES | 83.3 | 10.7 | 83.2 | 89.3 | 7.84 | 79.2 |

DDNR (n=35)

Cross-validation AUC: 0.77

Co-Morbid:

Myalgia: 65.7%

TMJ Arthralgia: 60.0%

|  | **Clinical Feature** | **Prevalence (%)** | **Non-Prevalence (%)** | **Sensitivity (%)** | **Specificity (%)** | **LR+** | **Importance Score** |
| --- | --- | --- | --- | --- | --- | --- | --- |
| 1 | Does the patient have limited opening that occurred suddenly and then the joint clicking stopped ? YES | 48.6 | 0.8 | 48.5 | 99.3 | 10.00 | 68.6 |
| 2 | Pattern - Continuous | 68.6 | 11.0 | 68.5 | 89.0 | 6.28 | 67.1 |
| 3 | Does patient report limited MAO jaw opening | 65.7 | 11.5 | 65.7 | 88.5 | 5.77 | 64.0 |

|  | Clinical<br>Feature | Prevalence (%) | Non-Prevalence (%) | Sensitivity (%) | Specificity (%) | LR+ | Importance Score |
| --- | --- | --- | --- | --- | --- | --- | --- |
|  | (<40)?<br>YES |  |  |  |  |  |  |

Tinnitus (ring) (n=26)

Cross-validation AUC: 0.96

Co-Morbid:

Myalgia: 73.1%

TMJ Arthralgia: 50.0%

|  | **Clinical Feature** | **Prevalence (%)** | **Non-Prevalence (%)** | **Sensitivity (%)** | **Specificity (%)** | **LR +** | **Importance Score** |
| --- | --- | --- | --- | --- | --- | --- | --- |
| 1 | Otologic Diagnoses | 100.0 | 2.1 | 100.0 | 97.9 | 10.00 | 98.7 |
| 2 | Does the patient have ringing in the ear? YES | 88.5 | 1.7 | 88.6 | 98.3 | 10.00 | 92.1 |
| 3 | Earache / Ear Problem (ringing, pulsing, stuffy, dizziness) | 92.3 | 6.5 | 92.4 | 93.5 | 10.00 | 91.5 |
| 4 | Please rate your ear problem (pain, ringing, stuffiness, hearing loss etc.) on average. [0=No problem, 10=Problem as bad as you can imagine] | 88.5 | 6.7 | 88.5 | 93.3 | 9.99 | 89.1 |
| 5 | Extraoral Ear | 76.9 | 3.7 | 76.8 | 96.3 | 10.00 | 83.8 |
| 6 | Pattern - Continuous | 69.2 | 3.0 | 69.3 | 97.0 | 10.00 | 79.8 |
| 7 | Cause - Unknown | 65.4 | 4.7 | 65.5 | 95.3 | 9.93 | 76.4 |
| 8 | Character - No pain | 53.8 | 1.9 | 53.9 | 98.1 | 10.00 | 71.2 |
| 9 | Onset - Years Ago | 53.8 | 2.6 | 54.0 | 97.4 | 9.99 | 70.8 |
| 10 | Side - Bilateral | 53.8 | 2.4 | 53.8 | 97.6 | 9.99 | 70.8 |

|  | Clinical<br>Feature | Prevalen<br>ce (%) | Non-Prevalen<br>ce (%) | Sensitiv<br>ity (%) | Specific<br>ity (%) | LR<br>+ | Importan<br>ce Score |
| --- | --- | --- | --- | --- | --- | --- | --- |
|  | y |  |  |  |  |  |  |

MYOFASCIAL PAIN (n=309)

Cross-validation AUC: 0.68

Co-Morbid:

Myalgia: 76.1%

TMJ Arthralgia: 74.8%

|  | **Clinical Feature** | **Prevalence (%)** | **Non-Prevalence (%)** | **Sensitivity (%)** | **Specificity (%)** | **LR+** | **Importance Score** |
| --- | --- | --- | --- | --- | --- | --- | --- |
| 1 | Active Trigger Points in jaw muscle (Taut Band with Referral Pain) on exam?<br>YES | 48.9 | 2.1 | 48.8 | 97.9 | 10.00 | 68.0 |

